# Systems Immunology Approaches to Understanding Immune Responses in Acute Infection of Yellow Fever Patients

**DOI:** 10.1101/2024.05.04.24306661

**Authors:** André N. A. Gonçalves, Priscilla R. Costa, Mateus V. Thomazella, Carolina A. Correia, Mariana P. Marmorato, Juliana Z. C. Dias, Cassia G. T. Silveira, Alvino Maestri, Natalia B. Cerqueira, Carlos H. V. Moreira, Renata Buccheri, Alvina C. Félix, Felipe M. Martins, Vanessa E. Maso, Frederico M. Ferreira, José D. A. Araújo, Amanda P. Vasconcelos, Patrícia Gonzalez-Dias, Rafick-Pierre Sékaly, Otavio Cabral-Marques, Jordana G. A. Coelho-dos-Reis, Daniela M. Ferreira, Esper G. Kallas, Helder I. Nakaya

## Abstract

In the 2018 yellow fever (YF) outbreak in Brazil, we generated new transcriptomic data and combined it with clinical and immunological data to decode the pathogenesis of YF. Our analysis of 79 patients highlighted distinct gene expression patterns between acute YF infections, other viral infections, and the milder infection induced by the live-attenuated YF-17D vaccine. We identified a critical role for low-density, immature neutrophils in severe outcomes, marked by the downregulation of genes such as PADI4, CSF3R, and ICAM1 in deceased patients. These genes are essential for neutrophil migration and maturation, suggesting their pivotal role in disease progression. Furthermore, our study revealed a complex interaction among inflammation-related genes: increased expression of CXCL10 in the acute phase was accompanied by decreased expression of IL-1b and an increase in IL1R2, a decoy receptor that binds to IL-1 to inhibit its activity. The diminished expression of HLA class II genes suggests an impairment in antigen presentation. These insights underscore the delicate balance of immune responses in YF pathogenesis and provide a foundation for future therapeutic and diagnostic advancements in managing YF.

**Highlights:**

● PBMC transcriptome analysis in acute YFV highlights different immune responses between survivors and deceased patients.
● Decreased expression of HLA class II genes in dendritic cells of deceased YFV patients indicates a critical impairment in antigen presentation and innate immunity.
● Elevated B cell activation markers, such as BLNK and TNFRSF13B, in fatal YFV cases suggest an overactive, potentially dysregulated B cell response.
● Increased expression of neutrophil-related genes, including DEFA1B and MMP9, in deceased YFV patients highlights neutrophils’ role in aggravating inflammation and tissue damage.

Graphical Abstract

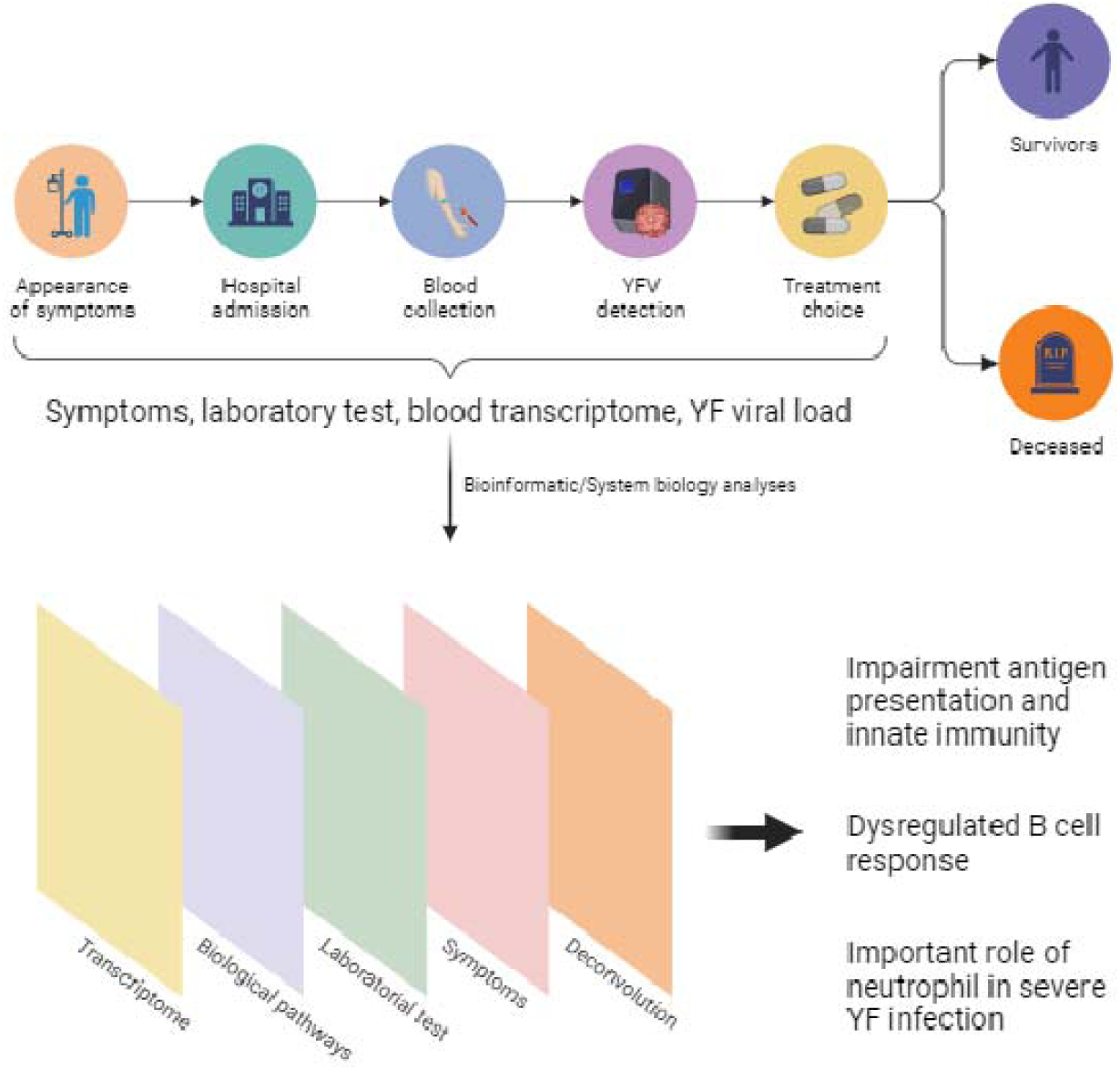

## Introduction

Yellow Fever (YF) is a significant public health concern in tropical regions of Africa and South America, accounting for an estimated 29,000 to 60,000 deaths annually (Garske et al. 2014; Paules and Fauci 2017; Song and Carneiro D’Albuquerque 2019). The disease can manifest as a spectrum of clinical presentations, from self-limiting, flu-like symptoms to severe forms characterized by hemorrhage and liver disease. Johansson et al. estimated that for every reported death, there are approximately 21 other infections, comprising 12 asymptomatic cases, 8 mild cases, and 1 severe but non-fatal case (Johansson, Vasconcelos, and Staples 2014).

The host immune response significantly influences disease severity. Studying immune modulators during early stages can identify markers linked to disease processes. These markers help pinpoint individuals at higher risk of severe outcomes (Fradico et al. 2023; van de Weg et al. 2023). The pathogenesis of YF involves various immune cells—including CD4^+^ T cells, CD8^+^ T cells, B cells, NK cells, macrophages, and antigen-presenting cells—alongside a complex interplay of pro-inflammatory and anti-inflammatory cytokines such as TNF, IFN-γ, and TGF-β (Pulendran 2009; Quaresma et al. 2013). Prior investigations have highlighted certain immune markers in response to Yellow Fever Virus (YFV) natural infection, such as increased hepatocyte apoptosis and elevated TNF-α and IFN-γ levels in the liver of fatal cases (Quaresma et al. 2013). During the Guinea epidemic, significantly higher serum levels of cytokines, including CXCL8, TNF-α, CCL2, IL-6, CXCL10, and IL-1Ra, were observed in fatal YF cases compared to survivors, a pattern echoed in subsequent studies (T. D. Querec et al. 2008; ter Meulen et al. 2004; Meier et al. 2009; Campi-Azevedo et al. 2012).

Our understanding of the global molecular mechanisms underlying the host immune response to YFV infection remains limited. This knowledge gap is primarily due to the reliance on systems vaccinology studies focusing on YF vaccines. These live-attenuated virus vaccine investigations have provided insights into general as well as YF-specific cytokines and cell types involved in the immune response to a mild infection. Studies have shown that vaccination with the YF vaccine can induce a polyfunctional human memory CD8^+^ T cell response, activate multiple Toll-like receptors on dendritic cells, and stimulate the production of a range of pro-inflammatory cytokines (Akondy et al. 2009; T. D. Querec et al. 2009; Pulendran 2009).

However, these findings, derived from a controlled vaccine response, may not fully capture the complexity of the immune response to natural wild-type YFV infection.

This study presents the first blood transcriptomic data from patients with acute YFV infection. We employed an integrative approach, combining molecular signatures with clinical and laboratory data. This strategy enabled a nuanced examination of immune response dynamics, distinguishing between the outcomes for survivors and those who succumbed to the disease. Our analysis connected gene expression patterns with severity markers, notably the Model for End-Stage Liver Disease (MELD) score, which evaluates liver function decline. We uncovered gene signatures indicative of YFV immunopathogenesis, such as the upregulation of genes driving the antiviral interferon response and plasma B cell differentiation and the downregulation of genes essential for antigen presentation in deceased patients. Moreover, our results suggest a critical role for low-density neutrophils in the pathogenesis of severe YF, underscoring their significance in combating YFV. Comparative analysis of the immune responses to the YF vaccine and other viral infections offered detailed insights into the distinctive immune mechanisms triggered by YFV natural infection. This work deepens our understanding of YF pathogenesis and opens pathways for developing targeted therapeutic interventions.

## Results

### Clinical Outcomes and Biochemical Markers in Yellow Fever Infection

We recruited 81 patients acutely infected with the YFV (Figure 1A). Among these patients, 26 died from the disease within the first 30 days of symptoms (32.9%), and 53 survived (67.1%). We excluded two patients (2.5%) who died after 30 days of post-onset symptoms from the analyses. Seventy-nine participants were included in the final analysis. Blood samples were collected during the acute phase (between 3 and 16 days after the onset of symptoms). Additionally, we collected blood samples from surviving patients during the convalescent phase (more than 30 days post-symptom onset), referred as “non-infected controls” (Figure 1A).

**Figure 1.**
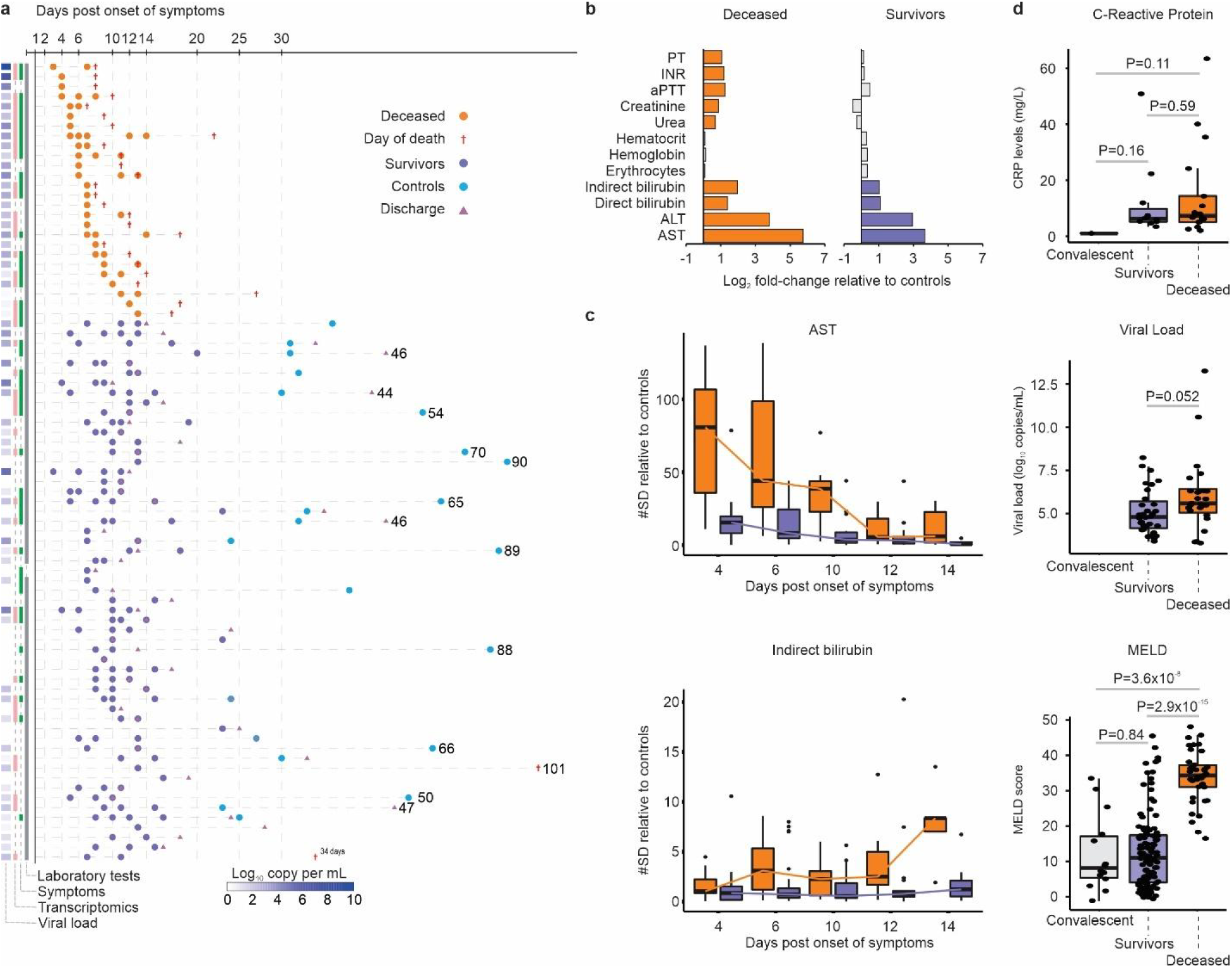
Patient Sample Distribution and Laboratory Indicators. (a) Distribution of deceased (orange dots) and survivors (purple dots) during the acute phase of YFV infection, indicating which assays were performed or data collected at each time point: transcriptome, symptoms, laboratory tests, and viral load measurement. The date of death is indicated by a red cross. Additional samples from surviving patients, collected post-recovery, are denoted as convalescent samples (blue dots), with a pink triangle indicating the hospital discharge date. Viral loads are represented by squares on the left, in copies per mL. (b) Fold-change in deceased and surviving patients’ laboratory parameter values relative to those in convalescent samples. Parameters are related to bleeding (PT [prothrombin time], INR [international normalized ratio], aPTT [activated partial thromboplastin time]), renal insufficiency (urea, creatinine), anemia (hematocrit, hemoglobin, erythrocytes), liver dysfunction (total bilirubin, indirect bilirubin, direct bilirubin), and hepatic injury (ALT [alanine transaminase], AST [aspartate aminotransferase]). (c) AST and indirect bilirubin levels for deceased and surviving patients compared with convalescent patients during the first 14 days post-symptom onset. (d) CRP levels, viral load, and MELD (Model for End-Stage Liver Disease) score for each patient outcome, for both non-infected and infected (deceased and survivor) patients. A nonparametric Wilcoxon test was applied to calculate differences between each group of samples.

Patients presented diverse symptoms; however, none demonstrated a statistical association with mortality (Figure S1). Analysis of blood parameters during the acute phase revealed pronounced liver injury (elevated levels of AST, ALT, indirect and direct bilirubin), renal impairment (increased creatinine and urea), and coagulopathy (increased PT, INR, and aPTT) compared to non-infected control patients (Figure 1B) (similar findings reported by (Kallas et al. 2019; Bailey et al. 2020)). Levels of AST were higher in deceased patients in the initial days post-onset of symptoms compared to survivors, while indirect bilirubin levels were higher in later days post-onset symptoms (Figure 1C). C-reactive protein (CRP) levels and YFV viral load did not significantly differ between survivors and deceased patients (Figure 1D). To assess the severity of these patients, we calculated a Model for End-Stage Liver Disease (MELD) score based on levels of bilirubin, INR, and creatinine (Malinchoc et al. 2000). As expected, deceased patients exhibited a higher MELD score compared to convalescent patients and survivors (Figure 1D).

### Dissecting the Transcriptional Landscape of YFV infection

To better understand the molecular mechanism of the disease, we first investigated the blood transcriptome of deceased and surviving patients during acute and convalescent phases. Initially, we calculated the molecular degree of perturbation (MDP score) of each sample (Gonçalves et al. 2019) to assess the molecular heterogeneity of the disease. At the transcriptome level, we found that deceased patients had higher MDP scores when compared to survivors (Figure S2A). Interestingly, the MDP score correlated with the MELD score, AST levels, and INR (Figure S2B-D), but not with viral load or CRP (data not shown).

Next, we identified gene signatures correlated with each one of the three factors associated with the immunopathogenesis of YFV infection analyzing the MDP scores of surviving and deceased patients. This information was taken from the Human Gene Atlas (Su et al. 2002) and we filtered each signature for highly expressed genes in six immune blood cell populations. As expected, transcription factors that mediate cellular responses to interferons, such as STAT1, STAT2, and IRF7, were associated with viral load across different cell types (Figure 2 and Figure S3). Interestingly, however, the MELD score was negatively correlated with HLA class II genes (HLA-DRA, HLA-DPA1, HLA-DMB, HLA-DPB1) in dendritic cells (Figure 2), suggesting an impairment in the innate immune system of patients with greater disease severity. Similarly, in CD8^+^ T cells, patients with a higher MELD score exhibited lower levels of the CCL5 gene (RANTES), which plays a crucial role in homing and migration of effector and memory T cells during acute infections, and ITGAL (CD11A), which potentially regulates effector CD8^+^ T cell activation and differentiation as well as antigen-specific central memory development in response to infection (Figure 2).

**Figure 2.**
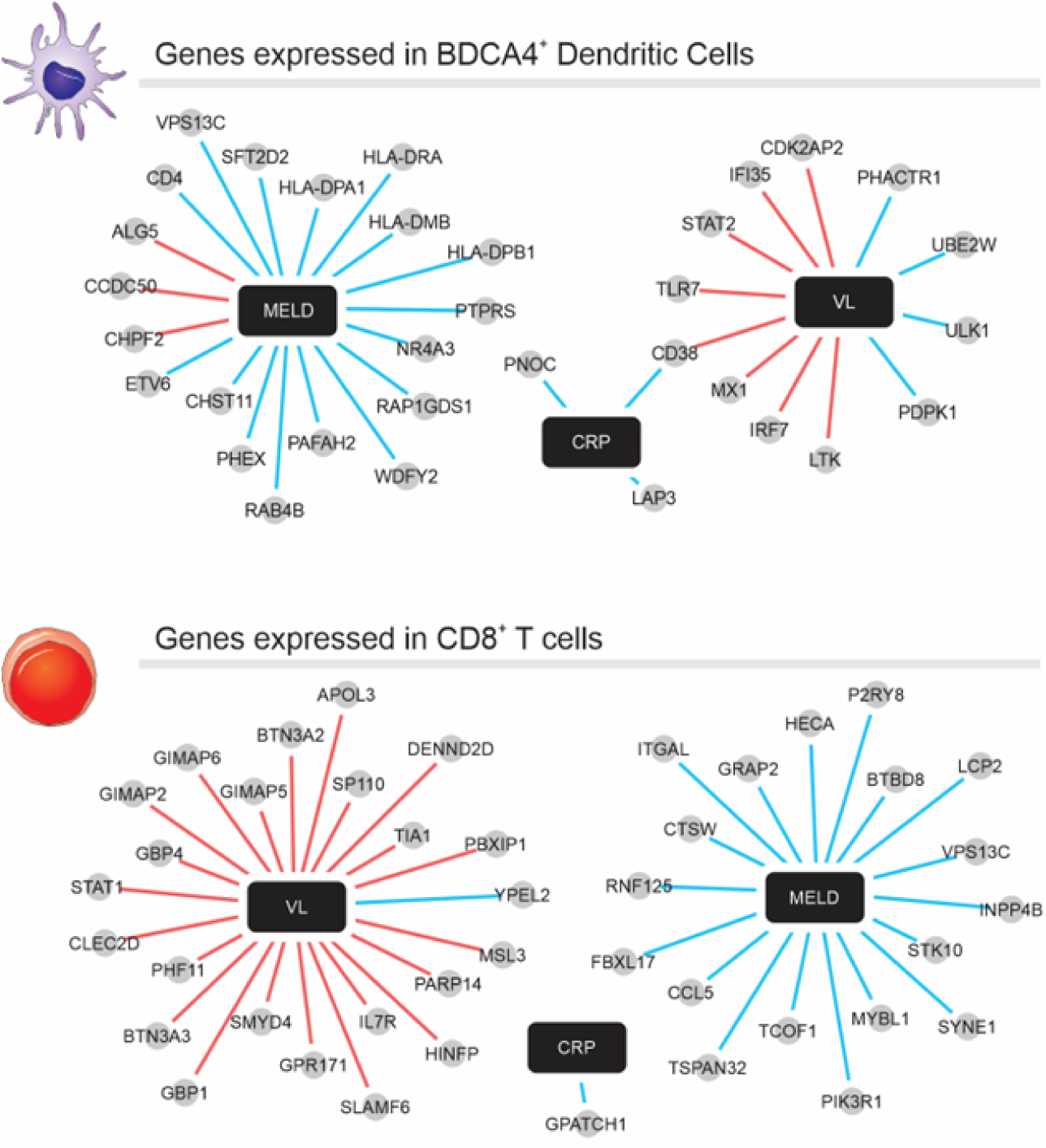
Clinical Parameter-associated Signatures in Plasmacytoid Dendritic Cells and CD8^+^ T Cells. The network was constructed using the correlation between gene expression (normalized to patients in the convalescent phase) and MELD score, C-reactive protein (CRP), or viral load (VL) values. Correlated genes (Spearman’s rank correlation coefficient > 0.5) that are also expressed in dendritic cells or CD8^+^ T cells according to the Human Gene Atlas were selected as nodes. The red line indicates a positive correlation, and the blue line indicates a negative correlation between genes (gray circles) and parameters (black rectangle).

We further detailed the impact of global expression changes caused by YFV infection, we performed a differential expression analysis between patients in the acute phase of the disease and those in the convalescent phase. We also compared the expression profile of patients who died from the disease to those who survived. We noticed that many genes were differentially expressed (DEGs) in both analyses (Figure 3A). Several genes encoding proteins related to the antiviral interferon response and the differentiation of B cells into antibody-secreting cells had increased expression in the acute phase of the disease and in patients who died from YF (Figure 3B and 3C). On the other hand, genes associated with antigen presentation, such as CD74 and several HLA class II genes, had decreased expression in patients who died from the disease (Figure 3C). Although the gene encoding the pro-inflammatory protein CXCL10 (IP10) was increased in the acute phase of the disease and associated with death, the gene encoding IL-1b had decreased expression (Figure 3A). The gene encoding the IL1R2 receptor, which binds to IL-1 and acts as a decoy receptor that inhibits the activity of IL-1 ligands, was also increased in the acute phase and in patients who died from YF. These data suggest that the role of inflammation and inflammasome-related pathways (or its suppression) during the first days of symptoms may be crucial to the outcome of the disease.

**Figure 3.**
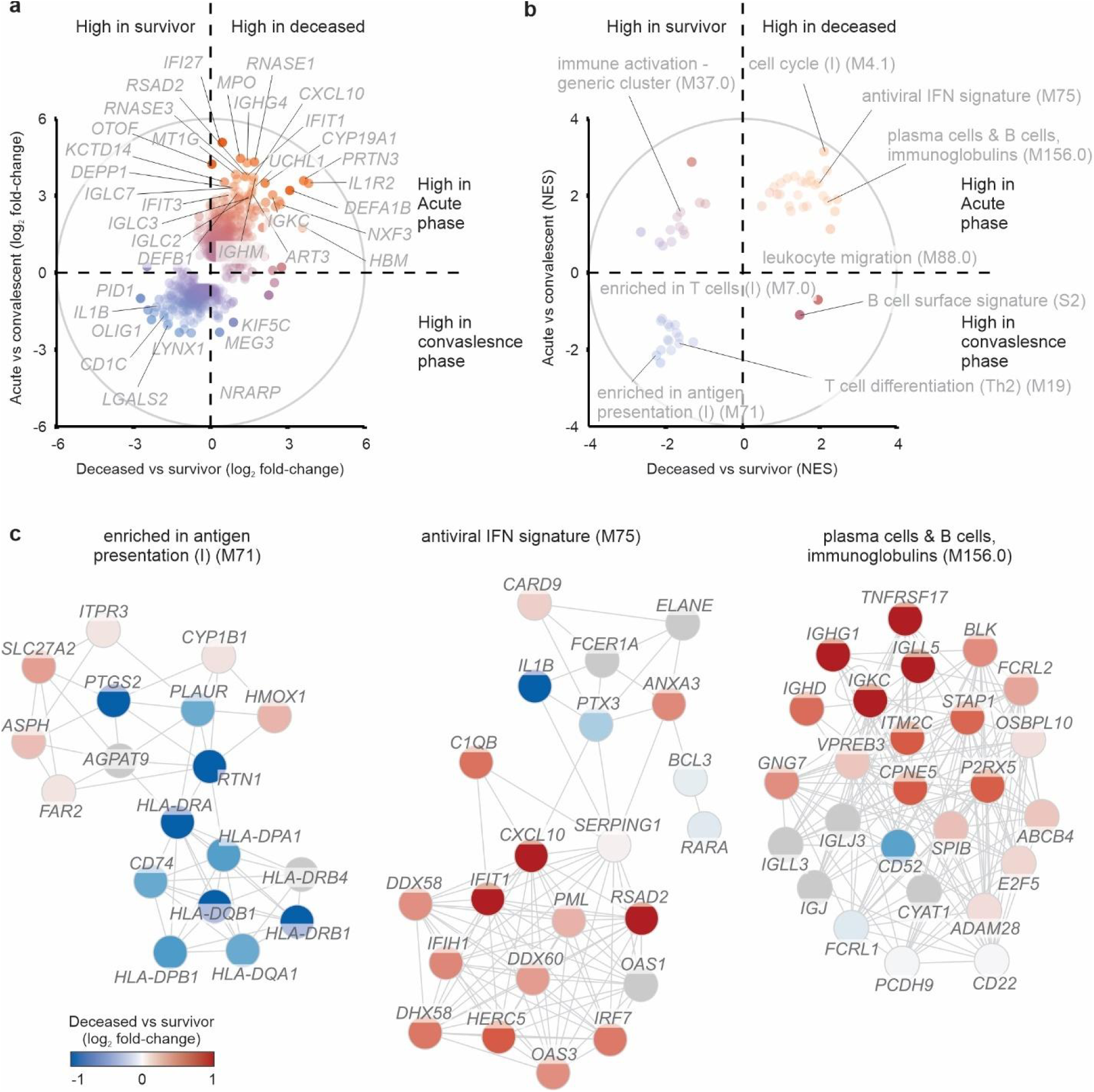
Transcriptomic Insights into Acute Infection and Disease Outcome. (a) Relationship between differentially expressed genes in deceased patients versus surviving patients and those during the acute phase versus the convalescent phase. Circles represent the log2 fold-change of genes that were up-regulated (red) or down-regulated (blue) in both comparisons. (b) Gene Set Enrichment Analysis (GSEA) using the log2 fold-change and adjusted p-value as rank and Blood Transcription Module (BTM) as gene sets. Circles represent the NES (Normalized Enrichment Score) of BTMs that were enriched with up-regulated (red) or down-regulated (blue) genes. (c) Network of antigen presentation (I) (M71), antiviral IFN signature (M75), and plasma cells and B cells, immunoglobulins (M156.0) gene sets. The edges of the BTM network were defined in our previous publication (S. Li et al. 2014).

### Potential Involvement of Low-Density Neutrophils in Yellow Fever Pathogenesis

Co-expressed genes are often functionally related or involved in the same biological process (Simone et al. 2021). We used CEMiTool to identify modules of genes with similar expression patterns (Russo et al. 2018). Among the 8 modules identified, we found that the activity of modules M5 and M6, which respectively contained genes related to plasma B cells and type I interferon response, was increased in deceased patients when compared to those who had already recovered from the disease (Figure S4A and S4B). In contrast, the activity of modules M2 and M3, which contained genes related to TLR signaling and inflammation, was decreased in patients who died from YF (Figure S4A and S4B). Within module M3 (Figure S4C), the TLR2 gene (which was not differentially expressed in any comparison) was identified as a hub in the co-expression network. It has been shown that the live-attenuated YFV strain of the YF-17D vaccine can activate Dendritic Cells through TLR2 to elicit the production of pro-inflammatory cytokines and IFN-a (T. Querec et al. 2006).

An interesting hub in module M2 was PADI4, which was also significantly down-regulated in deceased patients compared to survivors and convalescent patients (Figure S4C). PADI4 encodes one of the enzymes that convert arginine to citrulline residues. This protein promotes the decondensation of chromatin during the innate immune response to infection in neutrophils, being essential for the formation of neutrophil extracellular traps (NETs). Additionally, CSF3R and ICAM1 (are down-regulated in deceased patients compared to survivors) are also present in module M2. Both genes encode proteins essential for the adhesion, migration, and development of mature neutrophils (Yang et al. 2005; Dwivedi and Greis 2017). Although the role of neutrophils in severe YFV infection remains unclear (Fontoura, Rocha, and Marques 2021), our findings suggest that these cells play an essential role in the pathogenesis of the disease.

Our bulk RNA-seq data represents the transcriptome of a mixture of peripheral blood mononuclear cells (PBMCs). In the density gradient centrifugation method, high-density granulocytes (neutrophils, basophils, and eosinophils) are removed from the low-density fraction of PBMCs. However, low-density neutrophils, which are considered immature forms of neutrophils, can often be found in the PBMC fraction (Blanco-Camarillo, Alemán, and Rosales 2021) and are increased in certain pathological conditions such as sepsis, chronic infections, and autoimmune diseases (Schenz et al. 2021). In order to investigate the role of this type of neutrophil in the pathogenesis of the disease, we initially ran a deconvolution analysis using immature neutrophil signatures (Monaco et al. 2019) to estimate the relative frequency of this cell type in the PBMCs of YF patients. While 76% (16 out of 21) of YF patients who died from the disease had estimated immature neutrophil levels above 1%, this level was only seen in 50% (10 out of 20) of patients who survived (Figure 4A). Comparing the estimated levels of low-density neutrophils directly, we noticed a significant increase in patients who died compared to those who survived (Figure 4B). We then analyzed complete blood count data from patients and confirmed that the levels of immature neutrophils are associated with death from the disease, as well as the levels of mature neutrophils (Figure 4C, (Kallas et al. 2019)). Taken together, these results show for the first time that low-density neutrophils may have an essential role in driving NET production and inflammation in severe disease.

**Figure 4.**
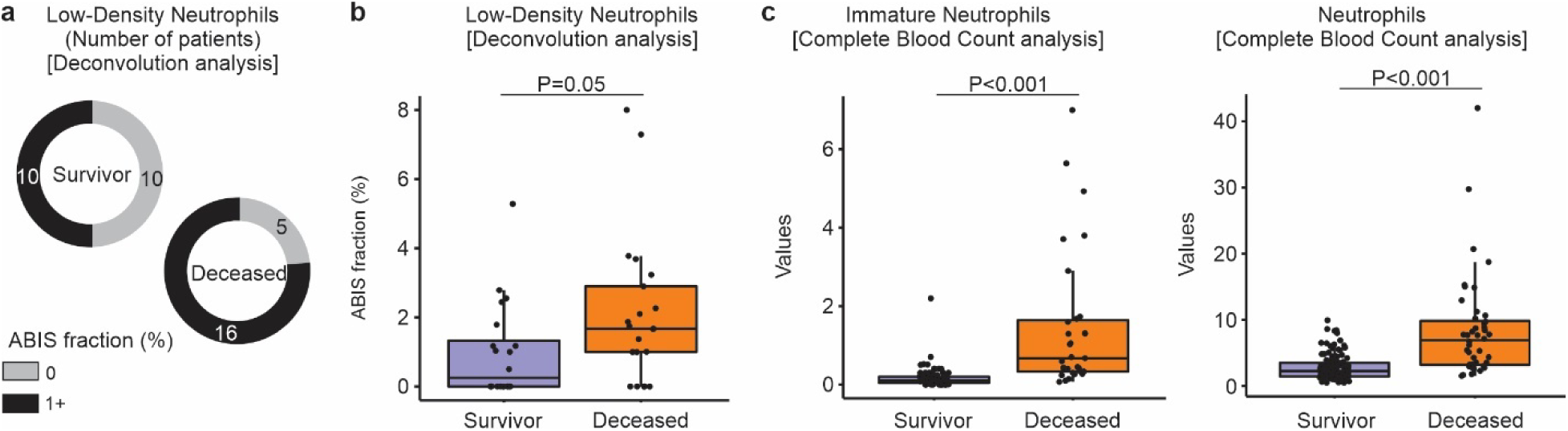
Levels of low-density neutrophils are increased in deceased YF patients. We used the ABsolute Immune Signal (ABIS) deconvolution tool (Monaco et al. 2019) to estimate the frequency of low-density neutrophils in the PBMC RNA-seq dataset. (a) Number of patients who survived or died from YF with ABIS fraction for low-density neutrophils over 1% (black). (b) ABIS fraction for low-density neutrophils estimated in patients who survived (purple) or died (orange) from the disease. (c) Number of immature neutrophils and neutrophils from Complete Blood Count (CBC) analysis of patients who survived (purple) or died (orange) from the disease. The Wilcoxon test was used to assess the statistical significance shown in each plot.

### Comparative Analysis of Transcriptome Changes in Acute YFV Infection and YF-17D Vaccination

Next, we compared the changes in the transcriptome resulting from acute infection with wild-type Yellow Fever Virus (YFV) to those from exposure to the live-attenuated YFV upon YF-17D vaccination. This involved re-analyzing microarray data from (Hou et al. 2017), which tracked the global expression profile of 21 volunteers given the YF-17D vaccine. The genes associated with the type I interferon response, previously described as genes induced post-vaccination (T. D. Querec et al. 2009), were similarly induced after acute infection (Figure 5A). In fact, of all genes with increased expression (2-fold or more) during acute infection compared to the convalescent phase, 242 (46.7%) were also induced after vaccination (Figure 4B). However, 276 genes did not exhibit increased expression post-vaccination, including *RNASE1*, *RNASE3*, *MPO*, *IL1R2*, *SERPINB2*, and *S100A12* (Figure 4B). Several of these genes, which are associated with modulation of inflammation (IL1R2, SERPINB2, and S100A12) or antiviral response (RNASE1, RNASE3, MPO), are expressed in monocytes and neutrophils. Enrichment analysis of signaling pathways suggested increased cell proliferation and B cell activity in infection and vaccination (Figure 5C). Yet, while pathways related to antigen presentation, immune response regulation, and T cell activation were triggered in the initial days post-vaccination, the activities of these pathways declined in acute infection or in patients who succumbed to the disease (Figure 5C). Thus, unlike infection with wild-type YFV, vaccination could effectively convey sustained immune response activation at the molecular level, in addition to connecting innate and adaptive immunity by stimulating antigen-presenting cells that in turn ultimately educate and trigger YFV antigen-specific T cells (Pulendran 2009).

**Figure 5.**
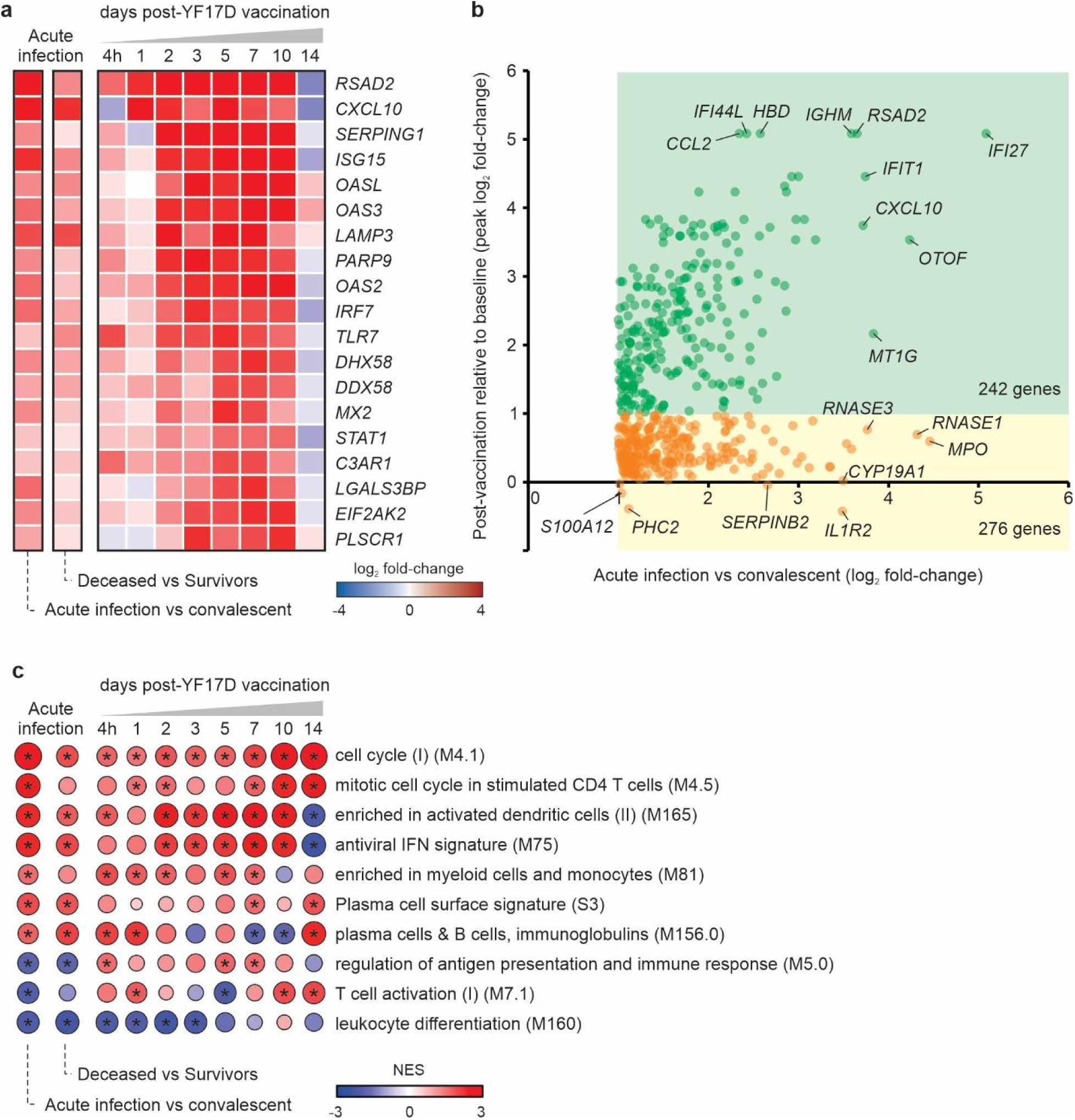
Comparison of acute YFV infection signatures and YF vaccination. (a) Type I interferon response-associated genes induced by the YF-17D vaccine as reported in (T. D. Querec et al. 2009). Genes are shown in rows and differential expression analyses in columns. The two left columns represent the comparison between patients in the acute phase of YFV infection compared to convalescent patients (first column) and between YF patients who died compared to patients who survived the disease (second column). The remaining columns show comparisons between the different timepoints (4h and 1, 2, 3, 5, 7, 10, and 14 days after vaccination) compared to the baseline of individuals vaccinated against YF. Colors represent the log2 fold-change of expression in each comparison. (b) Genes with increased expression in patients with acute YFV infection compared to convalescent patients (log2 fold-change > 1) that were or were not induced after YF vaccination. The values of log2 fold-change were re-scaled using the quantile. The x-axis represents the log2 fold-change values in acute infection and the y-axis represents the log2 fold-change values in vaccination (we used the value referring to the peak of induction in all time points). The green square shows genes that were also induced after vaccination (log2 fold-change > 1) and the yellow square shows genes that were not induced after vaccination. We indicated the total number of genes in the squares. We also indicated symbols of some selected genes. (c) Gene set enrichment analysis using log2 fold-change values as ranks and BTMs as gene sets. The selected BTMs are represented in rows and comparisons in columns. Colors represent the Normalized Enrichment Score (NES) and the asterisk indicates an adjusted P-value < 0.05.

### Comparative Analysis of Transcriptome Changes in Acute YFV Infection and Other Viral Infections

In addition, we investigated whether the signatures associated with death from YF could also be related to other viral infections. To do this, we compared the list of 276 genes with increased or decreased expression in the blood of YF patients who died from the disease compared to the convalescent phase with the differentially expressed genes in the blood of: (1) patients infected with the Dengue virus who had or did not have shock syndrome (1,341 genes, (Banerjee et al. 2017)); (2) patients infected with the Chikungunya virus compared with healthy controls (4,957 genes, (Soares-Schanoski et al. 2019)); and (3) patients with severe COVID-19 compared with healthy controls (704 genes, (Xiong et al. 2020)). Most of the genes (161 genes, 58.3%) in the YF signatures were not shared by infection with other viruses, suggesting that most genes are specifically associated with the YF pathophysiology. As expected, several genes related to the antiviral response had increased expression in YF and other viruses (Figure 6). Similarly, key genes related to inflammation such as PTGS2 (COX2), A2M, and KLRB1 or associated with immunity like MATK and PZP had decreased expression in patients who died from YF and in patients with COVID-19 or infected with CHIKV (Figure 6).

**Figure 6.**
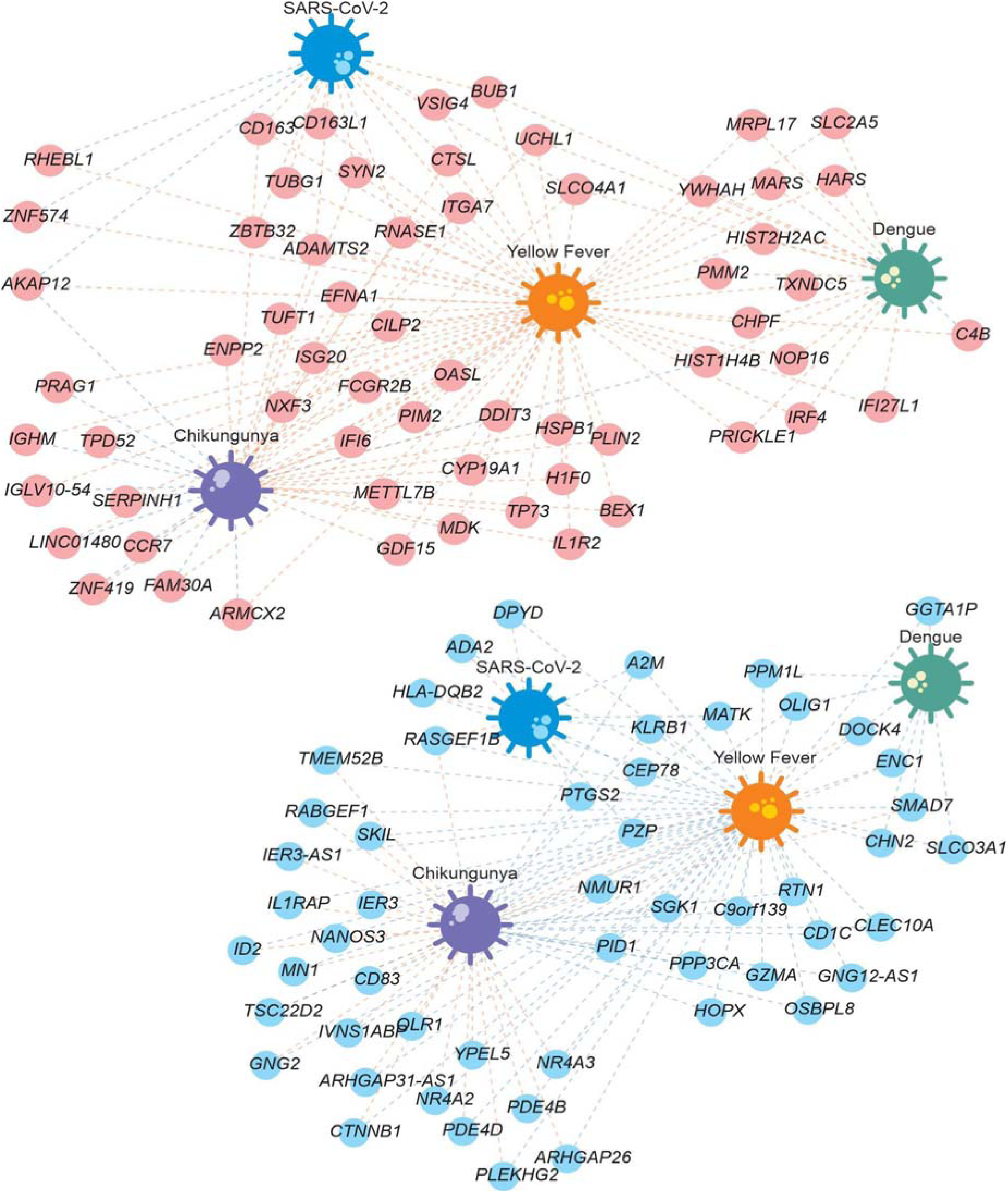
Comparing signatures associated with infection by Yellow Fever, Dengue, Chikungunya or SARS-CoV-2. The nodes represent the genes that were up-regulated (red nodes, top part) or down-regulated (blue nodes, bottom part) in patients who died of Yellow Fever compared with those who survived during the acute phase of the disease and who were also differentially expressed in infection by another virus. Red and blue edge colors indicate the genes up– or down-regulated by some virus, respectively.

Some genes had the opposite direction between the signatures of YF and other viruses. AKAP12 is a scaffold protein that is related to liver injury (Wu et al. 2022) and liver fibrosis (Lee et al. 2018) and showed increased expression in patients who died from YF but decreased in patients with COVID-19 or infected with CHIKV (Figure 6). Genes related to complement activation (C4B), lymphocyte homing (CCR7), or associated with immunoglobulins (FAM30A, IGHM, and IGLV10-54) (Lima et al. 2019) also had increased expression in patients who died from YF but decreased in patients infected with CHIKV (Figure 6). Among the genes with altered expression in patients who died from YF, several genes related to viral immunity had increased expression in other viruses, including RASGEF1B, ADA2, HLA-DQB2, DPYD, CD83, IL1RAP, and IVNS1ABP (Figure 6).

Our study identified unique alterations in the gene expression profiles of patients who succumbed to Yellow Fever (YF). Notably, two genes crucial for B cell development and activation – BLNK and TNFRSF13B (TACI) – showed exclusive changes in YF cases. We also observed a unique up-regulation of genes encoding hemoglobin components (HBA2, HBD, HBM) in those who died from YF. These genes may reflect precocious anemia that triggers innate immune responses by acting as damage-associated molecular patterns (Bozza and Jeney 2020). Additionally, we identified several genes exclusively up-regulated in YF cases that are key to neutrophil function. These include DEFA1B, a defensin released from specific granules upon activation to combat pathogens; MMP9, a protein involved in neutrophil migration by degrading extracellular matrix proteins; PRTN3, a serine protease that assists neutrophil migration by breaking down extracellular matrix components; and S100P, a protein that, upon release, acts as a signal to recruit and activate immune cells. Our analysis revealed unique and specific immune response pathways, both innate and adaptive, that may be associated with YF severity.

## Discussion

This comprehensive study investigated the clinical, biochemical, and transcriptional responses of YFV acute infection in 79 patients. Our findings have uncovered gene signatures that are associated with liver injury, renal impairment, and coagulopathy in patients during the acute phase. These findings highlight the potential novel role of low-density neutrophils in severe YFV cases. Additionally, our research compared these findings with the immune response to the YF-17D vaccine and acute infections with other viruses, providing valuable insights into the pathophysiology of the disease.

We connected the expression of genes such as STAT1, IRF7, PADI4, along with HLA-DRA, HLA-DPA1, and CCL5, to the severity of Yellow Fever Virus (YFV) infection, shedding light on the molecular underpinnings of the diverse clinical manifestations and complications associated with the disease and vaccinated individuals (T. D. Querec et al. 2008; Hou et al. 2017; Azamor et al. 2021). These genes, crucial for the immune response and inflammation regulation (Qing and Liu 2023; Saheb Sharif-Askari et al. 2021; Tolomeo, Cavalli, and Cascio 2022), provide a molecular basis for the observed biochemical markers of severe liver damage, renal impairment, and coagulopathy during the acute phase. This aligns with previous findings that have documented the profound effects of YFV on organ function (Bailey et al. 2020; Kallas et al. 2019; Z. Chen et al. 2016). The early elevation of AST levels (Z. Chen et al. 2016) and the subsequent rise in indirect bilirubin levels (Wouthuyzen-Bakker et al. 2017) in deceased patients indicate a pattern of organ damage closely related to the dysregulation of these genes.

Our study also identifies a potential impairment in the immune system’s antigen presentation capabilities, highlighted by the negative correlation between the MELD score and HLA class II gene expression in dendritic cells. That observation could indicate an impairment stemming from the viral modulation of host immune functions (Lin et al. 2022) or the inefficient presentation of viral antigens by certain HLA molecules (Augusto et al. 2023). Additionally, the observed decrease in genes crucial for T cell migration and memory, such as CCL5 and ITGAL, in cases with higher MELD scores (Lim et al. 2010) underscores disrupted immune processes critical for an effective response to infection. Combined with the clinical and biochemical markers of disease severity, these molecular insights compile a nuanced understanding of YFV’s pathogenic landscape. By integrating these observations, the utility of the MELD score as a prognostic indicator is reinforced, and a framework is also established for exploring the intricate virus-host interactions, offering potential therapeutic avenues to alleviate the burden of YFV infections.

Clinical inflammation during viral infections, including YFV (Olímpio et al. 2022; Casanova and Abel 2021), is characterized by significant leukocyte infiltration and activation within tissues, as well as an amplified activation of circulating leukocytes in response to viremia. This phenomenon indicates a dual failure: first, in the cell-intrinsic immunity ability to control the virus within leukocytes or other cells, and second, in the steady-state immune response capacity to curtail extracellular virus proliferation and infected cells (Casanova and Abel 2021). In the context of YFV infection, this complex interplay of inflammatory responses can either mitigate or exacerbate the disease progression, with the outcome varying according to individual patient responses and disease phases. Our findings demonstrate that in the case of YFV infection, expression of systemic inflammatory markers, such as IL-1b, IL-6, and TNF, are not increased in the acute phase compared to the convalescent phase. In fact, we observed that YFV patients who died from the disease compared to control individuals have lower expression levels of IL-1a, IL-1b, and IFN-g and significant elevation in the levels of IL1R2, a decoy receptor that binds to IL-1 to inhibit its action. Contrasting with these findings, a study (Pelletier et al. 2021) examining the cytokine storm in the same cohort of YFV infection identified a substantial upregulation of pro-inflammatory cytokines, including IFNα2a, IFN-β, IL-6, IL-18, and IFN-γ, in patients with severe disease outcomes. This cytokine storm, not directly correlated with viral load at hospitalization but potentially impeding viremia resolution, suggests that the inflammatory response magnitude and regulation, such as through IL1R2 elevation, play crucial roles in disease severity and outcomes (Pelletier et al. 2021). The discrepancy between our observations and the cytokine profiles reported highlights the complexity of the immune response to YFV infection, suggesting that the balance between pro-inflammatory and regulatory mechanisms is pivotal in determining the infection trajectory and clinical outcome. The variance between protein and transcript levels largely arises from the weak correlation of transcript expression with protein amounts for most genes (Bauernfeind and Babbitt 2017). Moreover, immune responses originating from tissue rather than blood cells (Farber 2021) might account for the differences in cytokine levels between protein and transcript data.

The previous findings suggest a paradox in which increased inflammatory gene expression does not necessarily lead to effective viral control or heightened immune response. Instead, the presence of IL1R2 might indicate a regulatory mechanism gone awry, in such a way that the immune system, despite being in a heightened state of alert, inadvertently suppresses key components of the inflammatory response crucial for controlling viral spread. This dual role of inflammation in YFV infection, where it both attempts to control the virus and simultaneously may dampen specific immune responses, highlights the complex nature of host-pathogen interactions and suggests that a balanced immune response is critical for survival. Therefore, the interplay between pro-inflammatory cytokines and regulatory mechanisms like IL1R2 could be pivotal in determining the disease outcome, emphasizing the need for further investigation into how inflammation can be modulated to improve therapeutic interventions against YFV.

Interleukin 27 (IL27) gene has been identified as a significant cytokine with increased expression in YF-infected patients compared to control groups. This cytokine is known for its intriguing role in immune regulation, particularly its potential anti-inflammatory effects on Th17 cells (Yoshida and Hunter 2015), which are a subset of pro-inflammatory T cells that produce interleukin 17 and can contribute to the pathogenesis of various infectious diseases (Wong et al. 2010; Fabbi, Carbotti, and Ferrini 2017). The up-regulated expression of this gene suggests that IL-27 may play a role in modulating the immune response against the YFV, possibly by inhibiting the inflammation caused by Th17 cells (Jouhault et al. 2023), which could be crucial in determining the severity of the disease. In addition, IL-27 signalling activates innate antiviral proteins and protects against flavivirus infection (Kwock et al. 2020). Further research could provide more insights into how IL-27 influences Th17 cell-mediated inflammation as well as the antiviral responses in YF.

Neutrophils mediate vascular inflammation through their dual role in response to either infection or an initial sterile injury, intricately interacting with platelets for bacteria trapping (Phillipson and Kubes 2011). Our study connects YFV pathogenesis to the increase of low-density neutrophils, a phenomenon commonly seen in conditions such as sepsis and chronic infections (Tay, Celhar, and Fairhurst 2020; Sun et al. 2022; Dumont et al. 2024). During acute inflammation, the body releases immature neutrophils, uniquely characterized by their potent antimicrobial abilities and partially segmented nuclei (van Grinsven et al. 2019). However, immature neutrophils amplify vascular inflammation in giant cell arteritis through their production of reactive oxygen species (ROS), exacerbating endothelial damage and contributing to the disease’s vascular complications (Wang et al. 2020). YF deceased patients exhibit an up-regulation of particular inflammation and vasculitis genes compared to survivors. Notably, genes like PRTN3 and MMP9 were highly expressed in deceased patients, potentially contributing to the severe vascular complications observed in fatal cases (Galis and Khatri 2002; Trivioli et al. 2022). In fact, immature neutrophils are a result of emergency granulopoiesis triggered by viral infections. The high demand of neutrophils during viral invasion imposes an increased turnover of these cells in the blood, which calls for an abundant efflux of newly differentiated granulocytes that may fail to maturate properly in the bone marrow. These recently egressed immature neutrophils in a higher proportion than usual display impaired functions and increased propensity to NET formation (Rawat, Vrati, and Banerjee 2021) as well as increased T cell suppression (Hassani et al. 2020). Therefore, immature neutrophils may activate pathways associated to a deleterious immune response in face of YFV infection.

Vaccination with YF-17D provides long-term protection and mimics a mild virus infection. YF-vaccinated individuals exhibit pronounced differences in gene expression compared to natural wild-type YFV infection. These differences can be attributed to the divergencies in the timeframe of the vaccination and the natural wild-type YFV exposure in which the days post-infection, also include the virus incubation period. The acute infections analyzed in this study, occurring on average 15 days post-symptoms onset and considering the incubation period of YFV (approximately 3 to 6 days, according to (WHO 2023)), indicate that natural infection has a more extended immune response time span. The differential gene expression observed post-vaccination highlights a rapid immune response, typically within 14 days. Importantly, it underscores the interplay between the innate and adaptive immune responses, which, in contrast to being compromised in natural infections, are fully functional in vaccinated subjects.

YFV is a member of the genus Orthoflavivirus (Postler et al. 2023) within the family Flaviviridae, which includes mosquito-transmitted viruses such as Dengue Fever Virus (DENV), Japanese Encephalitis, West Nile Virus, and Zika Virus (CDC 2022). Intriguingly, YFV-infected patients exhibited a unique signature of differentially expressed genes (DEGs) involved in B cell development and activation, as well as neutrophil functions, compared to those infected with other family members like DENV, unrelated mosquito-transmitted viruses such as Chikungunya Virus (CHIKV), and a respiratory virus (SARS-CoV-2). Additionally, genes associated with YFV pathogenesis, such as those linked to liver injury (Wu et al. 2022) and fibrosis (Lee et al. 2018), were identified. However, YFV displayed some biological pathways with responses opposite to those observed in other viruses. For example, complement activation, lymphocyte homing, and associations with immunoglobulins were increased, while several genes related to viral immunity decreased in YF cases. These findings suggest that YFV elicits a distinct immune response intricately connected to the severity of cases.

In this study, we acknowledge that our cohort is exclusively male, with an average age of 42 years (ranging from 18 to 72 years), which may limit the generalizability of our findings across genders and age groups. In fact, YF disease was significantly biased toward males as an occupational hazard, who were more susceptible to mosquito bites and viral exposure due to work in the field and open affected areas. The relatively small sample size, due to the availability of acute phase samples from severe YF patients, might reduce the statistical power of our analysis. However, we have applied multiple comparison corrections. Additionally, we were not able to collect and analyse samples from individuals without YFV exposure as endemic controls, which were gender and age-matched. We have instead selected the convalescent phase patients as a control to diminish inter-group genetic variability and focused solely on the transcriptome profile during the acute phase and after viral clearance. Furthermore, although our study used bulk sequencing to examine the immune response to YF, this method may not fully capture the heterogeneity of responses across different immune cell types. Single-cell sequencing, offering a detailed view of the immune landscape by revealing individual cell types of unique signatures and responses, represents a promising direction for future research. Therefore, our findings contribute essential insights into the interactions between YFV and the host immune system. However, further studies using single-cell sequencing are crucial to thoroughly understanding the complex cellular dynamics of YFV infection.

Taken together, our study presents a comprehensive integrative analysis, combining molecular data with clinical and laboratory parameters. Through this approach, we have identified novel molecules that potentially play a crucial role in acute YFV infection. These findings have the potential to serve as markers for new therapeutic targets or for evaluating the severity of the disease, vis-à-vis this life-threatening arboviral infection in future outbreaks.

## STAR METHODS

Detailed methods are provided in the online version of this paper and include the following:

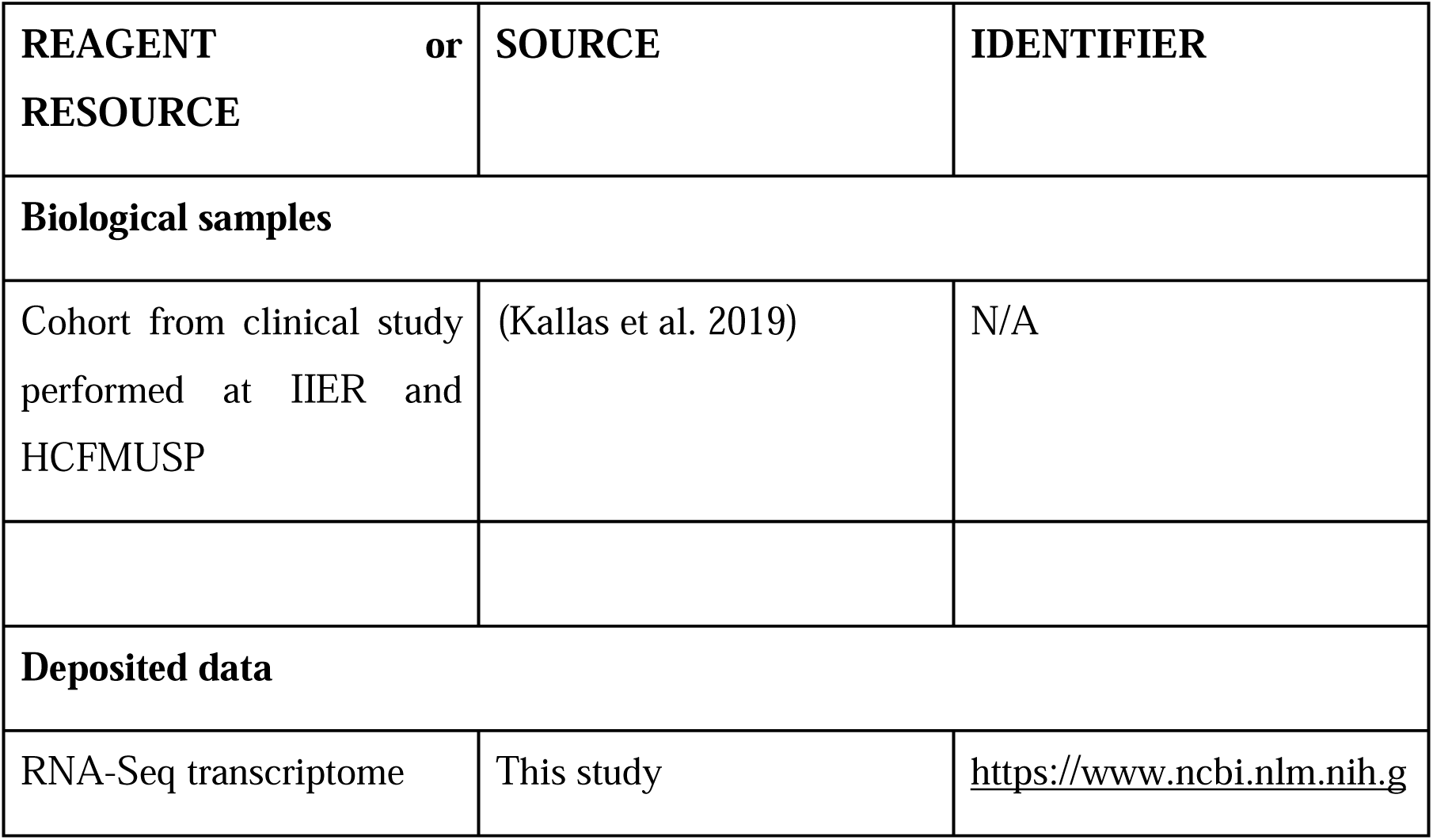
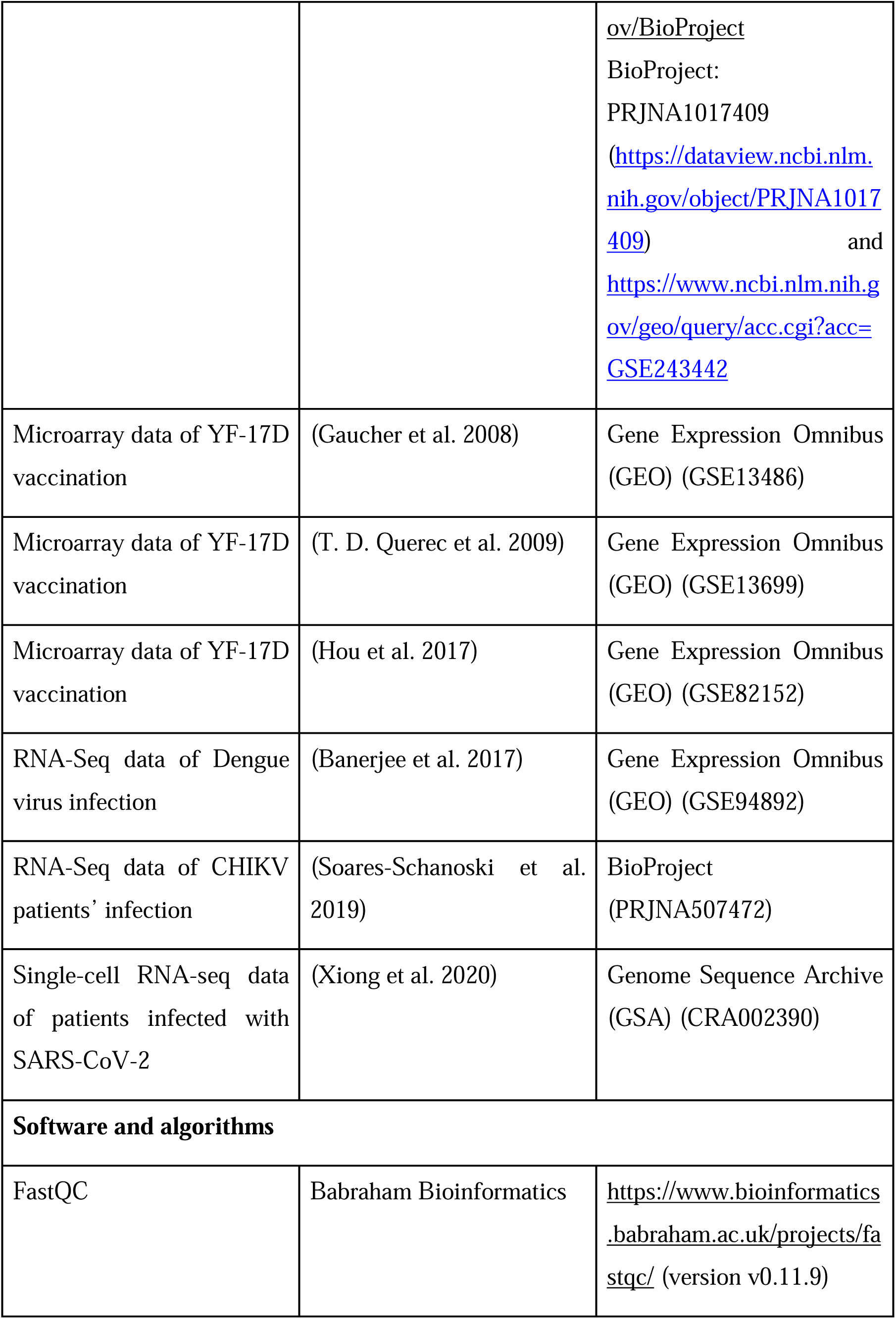
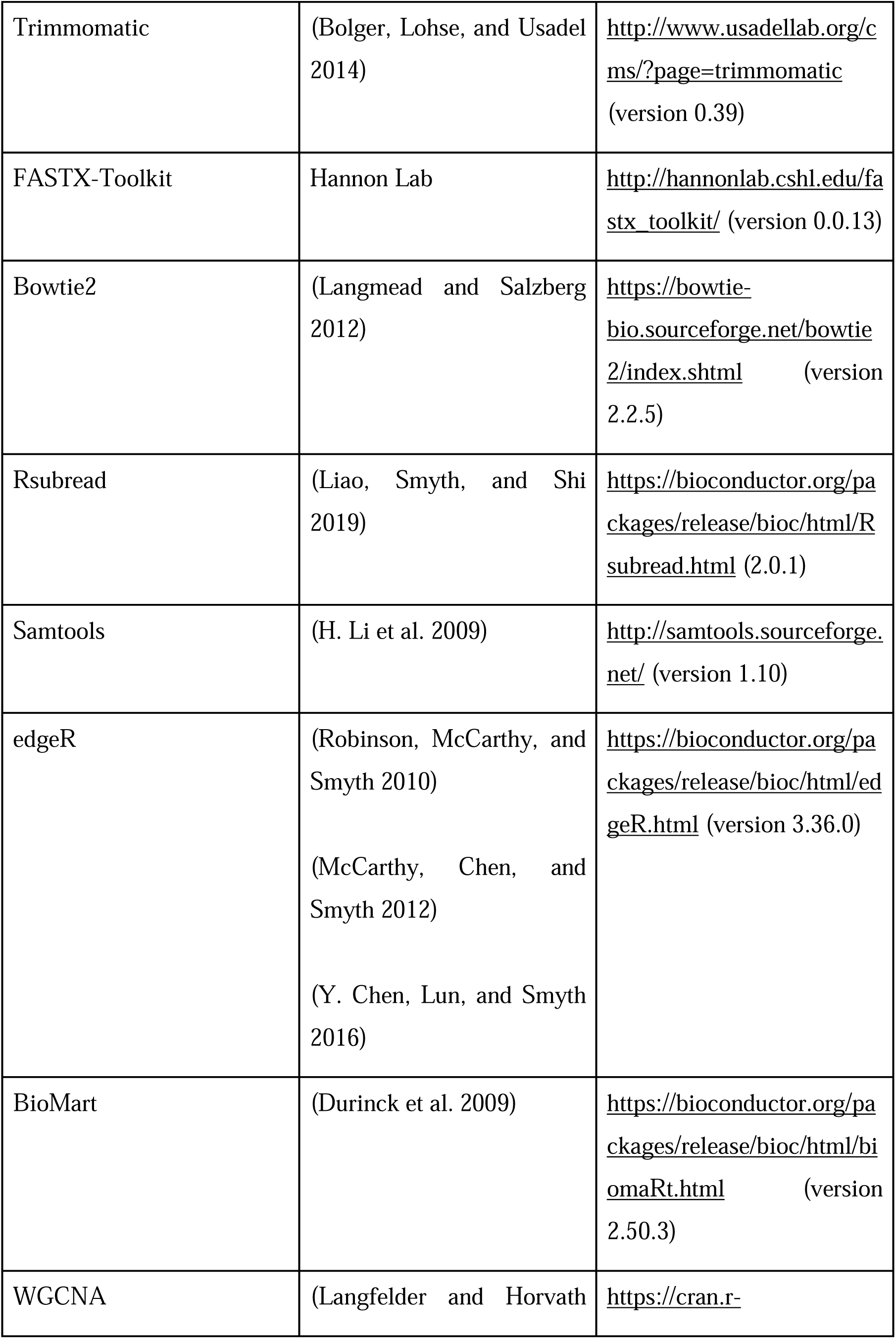
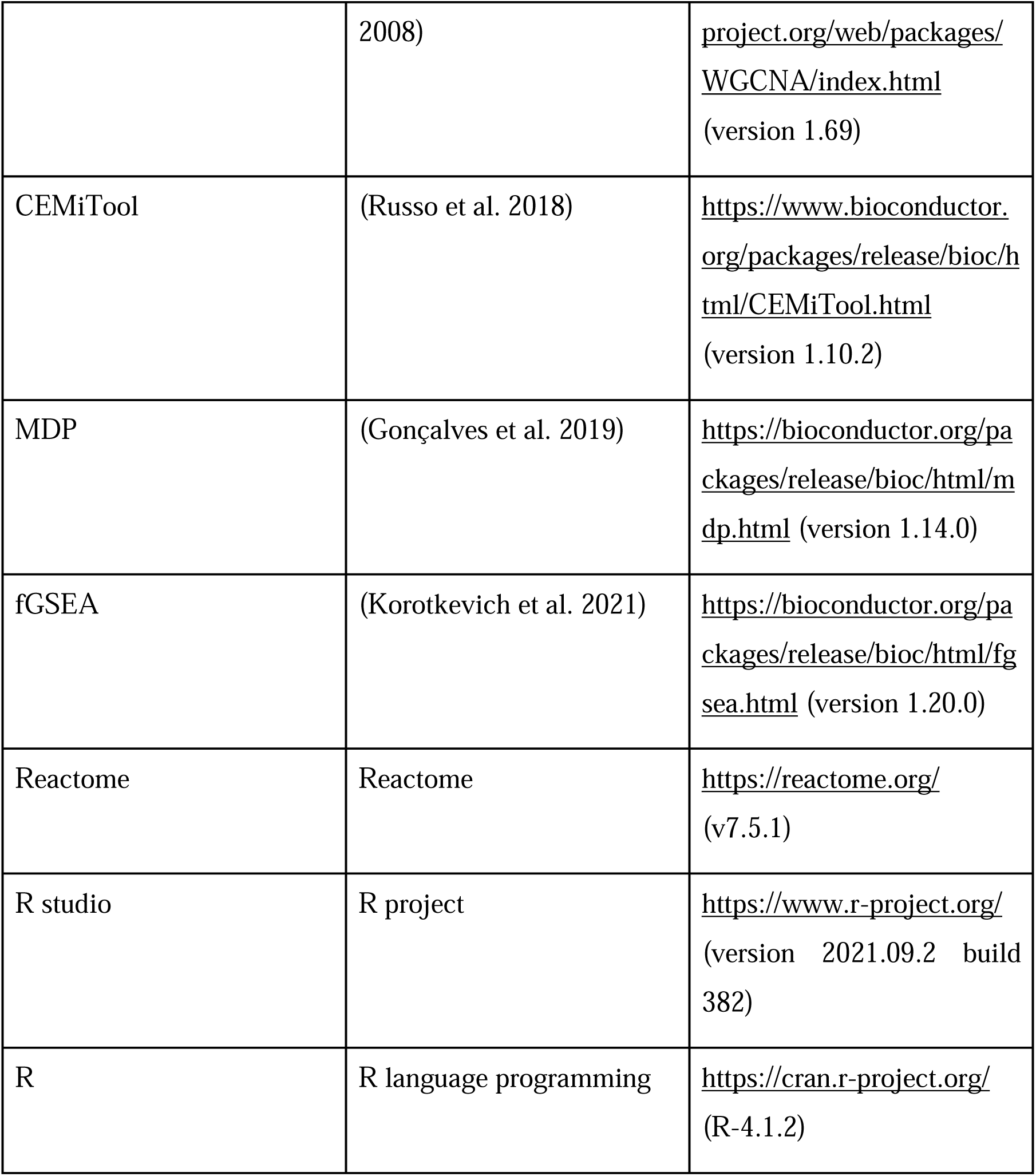
KEY RESOURCES TABLE.

## LEAD CONTACT

Further information and requests for resources and reagents should be directed to the lead contact, Esper G. Kallas or Helder I. Nakaya

## MATERIALS AVAILABILITY

This study did not generate new unique reagents.

## Experimental subject details

### Sample collection, ethical approval, and study design

On Jan 10, 2018, a referral system was established where patients with suspected yellow fever were admitted to one of two participating institutions: the Hospital das Clínicas, University of São Paulo, and the Infectious Diseases Institute “Emilio Ribas” (both located in São Paulo, Brazil). Patients older than 18 years admitted to the hospital with fever or myalgia, headache, arthralgia, oedema, rash, or conjunctivitis were consecutively screened for inclusion in the present study. The study received ethical approval from the ethical review boards at the Institute of Infectology “Emilio Ribas” and Hospital das Clinicas, University of São Paulo (CAPPesq: 15477; CAAE: 59542216.3.1001.0068). All patients provided consent for participation, or their legal representatives did so when required, before being included in the study. Confidentiality of patient identifiable information was strictly maintained during the study. Consenting patients were included if they had travelled to geographical areas where YFV cases had been previously confirmed. Patients were confirmed to be infected by real-time PCR by detecting the YFV RNA in blood samples collected at hospital admission or tissues at the autopsy. Suspected cases that tested negative for YFV RNA in blood had their diagnosis confirmed when tissue was positive for YFV RNA and presented pathological findings compatible with the disease. All patients were followed until death or for 60 days after enrollment, whichever occurred first. Peripheral blood mononuclear cells (PBMCs) were obtained by a Ficoll-Paque density gradient technique (Sigma Aldrich, St Louis), frozen, and stored at liquid nitrogen until use. The samples collected at admission (acute phase of infection) and more than 30 days post onset symptoms (convalescent phase) were submitted to load RNA virus (as described by (Kallas et al. 2019)) and RNA sequencing.

### Clinical data and standard laboratory test

The clinical data and standard laboratory tests were collected and analyzed at clinical laboratories located at the Hospital das Clínicas, School of Medicine, University of São Paulo, and the Infectious Diseases Institute “Emilio Ribas”. Demographic and clinical data were age, sex, race, duration of symptoms upon admission, oliguria, bleeding, lethargy, somnolence, muscle pain, nausea, abdominal pain, fever, headache, jaundice, coluria, vomiting, pain in joints, chills, diarrhea, pain behind the eyes, cough, arthritis, shaking, dyspnea, itching, sore throat, rash, coryza, conjunctivitis, and shock. The following laboratory tests were done at admission: prothrombin time (PT), international normalised ratio (INR), activated partial thromboplastin time (aPTT), creatinine, urea, hematocrit, haemoglobin, erythrocytes, indirect bilirubin, direct bilirubin, aspartate transaminase (AST), alanine aminotransferase (ALT), and c-reactive protein (CRP).

### Model for End-Stage Liver Disease (MELD) Score

The MELD score was determined using bilirubin, INR, and creatinine values for each patient. The formula used for the score was MELD = 3.78 x ln(bilirubin) + 11.2 x ln(INR) + 9.57 x ln(creatinine) + 6.43 (Singal and Kamath 2013).

### Bulk samples preparation and RNA sequencing

RNA from PBMCs was extracted using the RNeasy Mini Kit (Qiagen, Hilden, Germany), quantified using Nanodrop ND-1000 Spectrophotometer (Thermo Fisher Scientific), and integrity checked in a 2200 TapeStation system using RNA ScreenTape (Agilent Technologies, Santa Clara, CA). The cDNA libraries were constructed using the QuantSeq 3’ mRNA-Seq Library Prep Kit for Illumina FWD (Lexogen GmbH, Austria), following the manufacturer’s protocol. The concentration and the median size of the libraries were assessed by 2200 TapeStation with a DNA1000 ScreenTape (Agilent). The final pool of libraries was quantified by qPCR using the Kapa Sybr Green qPCR Kit (Roche Diagnostics, Mannheim, Germany) and subjected to a single-end sequencing (75 bp) in a NextSeq 500 Sequencing with NextSeq 500/550 High Output Kit v2.5 (Illumina, San Diego, CA).

### Quality control, mapping, and annotation processes

Raw single-end reads were preprocessed for quality control. Sequencing quality was assessed before and after adapter trimming using the program FastQC (Andrews 2010). The Trimmomatic software version 0.39 (Bolger, Lohse, and Usadel 2014) was used to remove adapters sequence trimming the 5′, and 3′ ends with a mean quality score below 25 (Phred+33) and discard reads shorter than 36 bp after trimming. We used the parameters “LEADING:3 TRAILING:3 SLIDINGWINDOW:4:20 MINLEN:36”. After preprocessing, the high-quality reads were mapped into the reference genome Homo sapiens (version GRCh38) with the bowtie2 program (version 2.2.5) (Langmead and Salzberg 2012).

To quantify the gene abundance of the mapped reads in each sample, we used the featureCounts tool from the R/Bioconductor package Rsubread (Liao, Smyth, and Shi 2019) with the following parameters: GTF.featureType = “gene”, GTF.attrType = “gene_id”, isPairedEnd = FALSE, minOverlap = value 1, allowMultiOverlap = FALSE, countMultiMappingReads = FALSE. Normalization of the gene counts was performed with counts per million normalization (CPM) and quantile normalization, which accounts for differences in library size.

### Molecular degree of perturbation and gene co-expression module

Sample quality and the molecular degree of perturbation (MDP) between groups were assessed with the R/Bioconductor package MDP (Gonçalves et al. 2019), using the normalized gene expression for survivor and non-survivor at the acute phase, which was compared to the convalescent group.

### Spearman correlation

We used the Spearman method to determine the correlation between two variables, considering p-value < 0.05 as correlated pairs.

### Differentially Expressed Genes Analysis

The identification of differentially expressed genes (DEGs) between groups was carried out with the R/Bioconductor package EdgeR (Robinson, McCarthy, and Smyth 2010; McCarthy, Chen, and Smyth 2012), using independent 2-class t-tests. P values were submitted to a false-discovery rate (Benjamini-Hochberg procedure) correction, and the statistical significance was defined by the criteria, adjusted p-value ≤ 0.05 and fold-change > 2.

### Pathway Enrichment Analysis

Regarding the pathways that may be related to the progression of the disease, a Gene Set Enrichment Analysis (GSEA) was performed using as ranks of the acute vs. convalescent and deceased vs. survivor comparison. A set of Blood Transcriptional Modules (BTM), previously identified by our group (S. Li et al. 2014) through large-scale network integration of publicly available human blood transcriptome, were used as the gene sets.

### Co-expression analysis

We identified and analyzed co-expression modules using the R/Bioconductor package CEMiTool (Russo et al. 2018). We used the log2CPM normalized gene expression matrix to identify the gene co-expression modules. The CEMiTool default parameters were followed. The biological meaning of the co-expression modules was explored by performing additional analysis with CEMiTool, the GSEA, the Over-Representation Analysis (ORA), and the network analysis. The GSEA was conducted to identify the induced and repressed modules in the three phenotypes: 1) patients in the acute phase deceased; 2) patients in the acute phase survived; and 3) convalescent patients. To identify the gene sets enriched in each module, we performed an ORA providing the Blood Transcriptional Modules (BTM) (S. Li et al. 2014) as gene sets. We provided Protein-Protein Interaction (PPI) data as a gene interactions file to build the network.

### Cell proportion estimation

The cell proportion was estimated using the ABsolute Immune Signal (ABIS) deconvolution tool (Monaco et al. 2019) and 29 human immune cell types were characterized by RNA-seq and flow cytometry.

## Microarray from published vaccination data

### Obtaining data from Gene Expression Omnibus repository

We used microarray data from three published data (Gaucher et al. 2008; T. D. Querec et al. 2009; Hou et al. 2017), identified under GEO id numbers GSE13486, GSE13699, and GSE82152, respectively. The time points combining all three studies comprehend in 4 hours, 1, 2, 3, 5, 7, 10 14 days post-YF-17D vaccination. The raw data expression and phenotypic data were obtained from the GEO database (https://www.ncbi.nlm.nih.gov/geo/) using getGEO, pData, and GSEMatrix functions from the R/Bioconductor package GEOquery.

### Quality control and preprocessing microarray data

Raw data from microarray datasets were submitted to the R/Bioconductor package arrayQualityMetrics for aberrant sample detection (Kauffmann, Gentleman, and Huber 2008). Samples flagged as outliers by two of the three tests performed by the package were removed. The resulting set of samples was iteratively resubmitted to arrayQualityMetrics using the same criteria until no samples were removed. Microarray datasets were then log-transformed and quantile normalized. Samples from Illumina platforms were normalized using functions provided by the R/Bioconductor package limma (Ritchie et al. 2015), whereas samples from Affymetrix platforms were normalized using Robust Multi-Array Average (RMA) from the R/Bioconductor package affy (Bolstad et al. 2004). Probes targeting genes in common were summarized by selecting the ones showing the highest mean of expression between all samples. Genes whose mean of expression was in the first decile were removed.

### Differentially expressed gene (DEG) analysis

The DEG analysis was obtained using the R/Bioconductor package limma, normalized expression, and phenotypic data for each timepoint post-YF-17D vaccination relative to samples before vaccination (D0). The lmFit() function was used to fit a linear model to the data accounting for the batch effects. To calculate t-statistics and P values for each gene we used eBayes function. Finally, we used the topTable() function to generate a table (containing information about the gene ID, expression values, log fold change, and p-value or FDR) for all genes detected in all studies and then used it for downstream analysis.

### Comparison between bulk transcriptome and microarray data

To compare vaccination and natural infection data, we normalized log2 fold-change values using a modified quantile using convalescent-phase samples from natural infection as reference group samples.

### Virus-related published infection studies

For the Dengue virus infection, we obtained differentially expressed gene tables from GEO id number GSE94892 processed by (Banerjee et al. 2017). Peripheral blood mononuclear cells (PBMCs) in clinically and virologically well-characterized patients with mild and severe dengue infection were used for Illumina HiSeq 2500 sequencing. The authors defined the group of samples A, B, and C, corresponding to Dengue Fever, Dengue Haemorrhagic Fever, and Dengue Shock Syndrome, respectively. The DEGs were identified with an adjusted p-value < 0.05 and fold-change > 2.

We analyzed and published a bulk RNA sequencing for CHIKV patients’ infection, and we used them (Bioproject number PRJNA507472) for viruses’ comparison. Blood samples were collected from subjects reporting arbovirus-like symptoms, and total RNA was extracted and sequenced using the HiSeq Illumina platform. The gold standard bulk sequencing pipeline was used as described in the publication (Soares-Schanoski et al. 2019). The DEGs were identified with an adjusted p-value < 0.05 and fold-change > 2.

The COVID-19 study was obtained from (Xiong et al. 2020) under the accession number CRA002390. The DEGs were provided in the supplementary data from the author’s paper. Peripheral blood mononuclear cells in COVID-19 patients were sequenced in the MGISEQ-2000 platform (MGI, Shenzhen, P. R. China). The DEGs followed the criteria adjusted p-value < 0.05 and fold-change > 2.

## Data and code availability

The complete set of raw sequences for each assay generated in the HiSeq Illumina 1500 were deposited at the NCBI through the BioProject: PRJNA1017409 and the BioSample Range SAMN37395281 to SAMN37395335.

## Data Availability

All data generated in the current study are deposited at https://www.ncbi.nlm.nih.gov/BioProject (BioProject: PRJNA1017409) and will be made publicly accessible upon publication of the article in a peer-reviewed journal.

https://dataview.ncbi.nlm.nih.gov/object/PRJNA1017409

https://www.ncbi.nlm.nih.gov/geo/query/acc.cgi?acc=GSE243442

https://yellowfeverdb.sysbio.tools

## Acknowledgements

We remember and honour the significant contributions of Luiz Gonzaga Francisco de Assis Barros D’Elia Zanella (*in memoriam*), whose wisdom and dedication greatly enriched this work. His passion for science and commitment to discovery continue to inspire us.

This study was supported by Bill & Melinda Gates Foundation, FAPESP, HCFMUSP, and USP. HIN is funded by FAPESP (grant numbers: #2017/50137-3, 2012/19278-6, 2018/14933-2, 2018/21934-5, and 2013/08216-2) and CNPq (313662/2017-7).

We are particularly thankful to the following grants for fellows: ANAG (FAPESP Process 2019/13880-5), VEM (FAPESP Process 2019/16418-0), APV (FAPESP Process 2019/27146-1), MVT (FAPESP Process 2019/13713-1 and 2016/01735-2) and the Coordenação de Aperfeiçoamento de Pessoal de Nível Superior (CAPES) (PhD Scholarships to CAC and MPM).

## Author contributions

Conceptualization, A.N.A.G., P.R.C., E.G.K., and H.I.N.; Data curation, A.N.A.G.; Formal Analysis, A.N.A.G., F.M.M., V.E.M., F.M.F., J.D.A.A., A.P.V., P.C.G.D.C., and H.I.N.; Funding acquisition, E.G.K., and H.I.N.; Investigation, A.N.A.G., R.P.S., C.O.M., J.G.A.C.R., D.M.F., E.G.K., and H.I.N.; Methodology, A.N.A.G., P.R.C., M.V.T., C.A.C., M.P.M., J.Z.C.D., C.G.T.S., L.G.F.A.B.E.Z., A.M., N.B.C., C.H.V.M., R.B., A.C.F., V.E.M., F.M.F., J.D.A.A., A.P.V., P.C.G.D.C., E.G.K., and H.I.N.; Project administration, E.G.K.; Resources, A.N.A.G., P.R.C., M.V.T., C.A.C., M.P.M., J.Z.C.D., C.G.T.S., L.G.F.A.B.E.Z., A.M., N.B.C., C.H.V.M., R.B., A.C.F., E.G.K., and H.I.N.; Software, A.N.A.G., and H.I.N.; Supervision, E.G.K., and H.I.N.; Validation, A.N.A.G., P.R.C., M.V.T., C.A.C., M.P.M., J.Z.C.D., C.G.T.S., L.G.F.A.B.E.Z., A.M., N.B.C., C.H.V.M., R.B., A.C.F., V.E.M., and H.I.N.; Visualization, A.N.A.G., V.E.M., F.M.F., J.D.A.A., A.P.V., P.C.G.D.C., and H.I.N.; Writing – original draft, A.N.A.G., F.M.M., and H.I.N.; Writing – review & editing, A.N.A.G., P.R.C., M.V.T., C.A.C., M.P.M., J.Z.C.D., C.G.T.S., L.G.F.A.B.E.Z., A.M., N.B.C., C.H.V.M., R.B., A.C.F., F.M.M., V.E.M., F.M.F., J.D.A.A., A.P.V., P.C.G.D.C., R.P.S., C.O.M., J.G.A.C.R., D.M.F., E.G.K., and H.I.N.

## Supplementary figures

**Figure S1.**
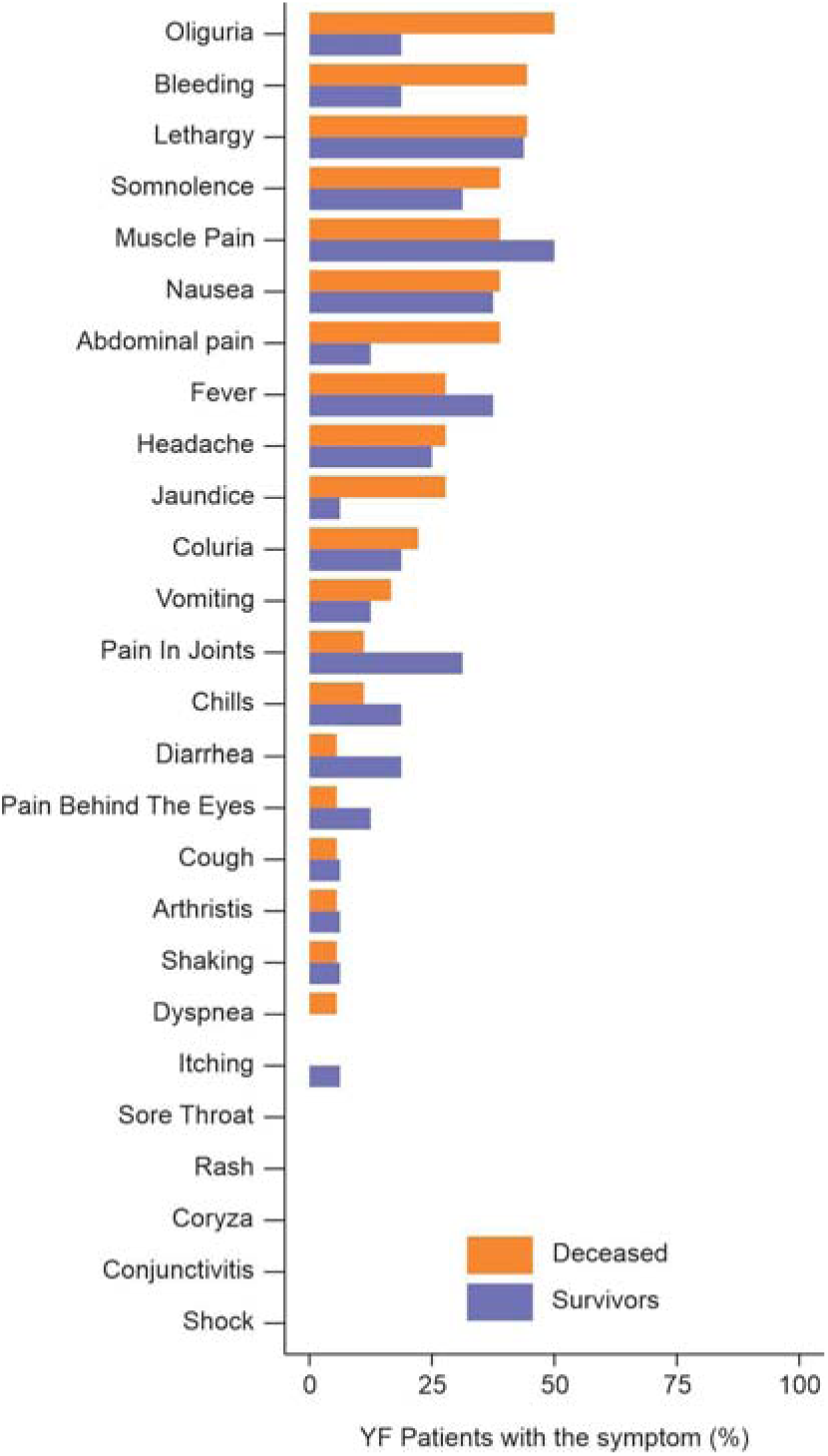
Summary of Symptoms in Yellow Fever Patients. The prevalence of each symptom among Yellow Fever (YF) patients was calculated as the frequency of occurrence relative to the total patient cohort. Fisher’s exact test was employed to determine the prevalence disparities of symptoms between deceased patients and survivors. Additionally, a proportion test was utilized to evaluate the differences in symptom prevalence among all YF patients. No statistically significant differences were observed for any of the symptoms when comparing outcomes.

**Figure S2.**
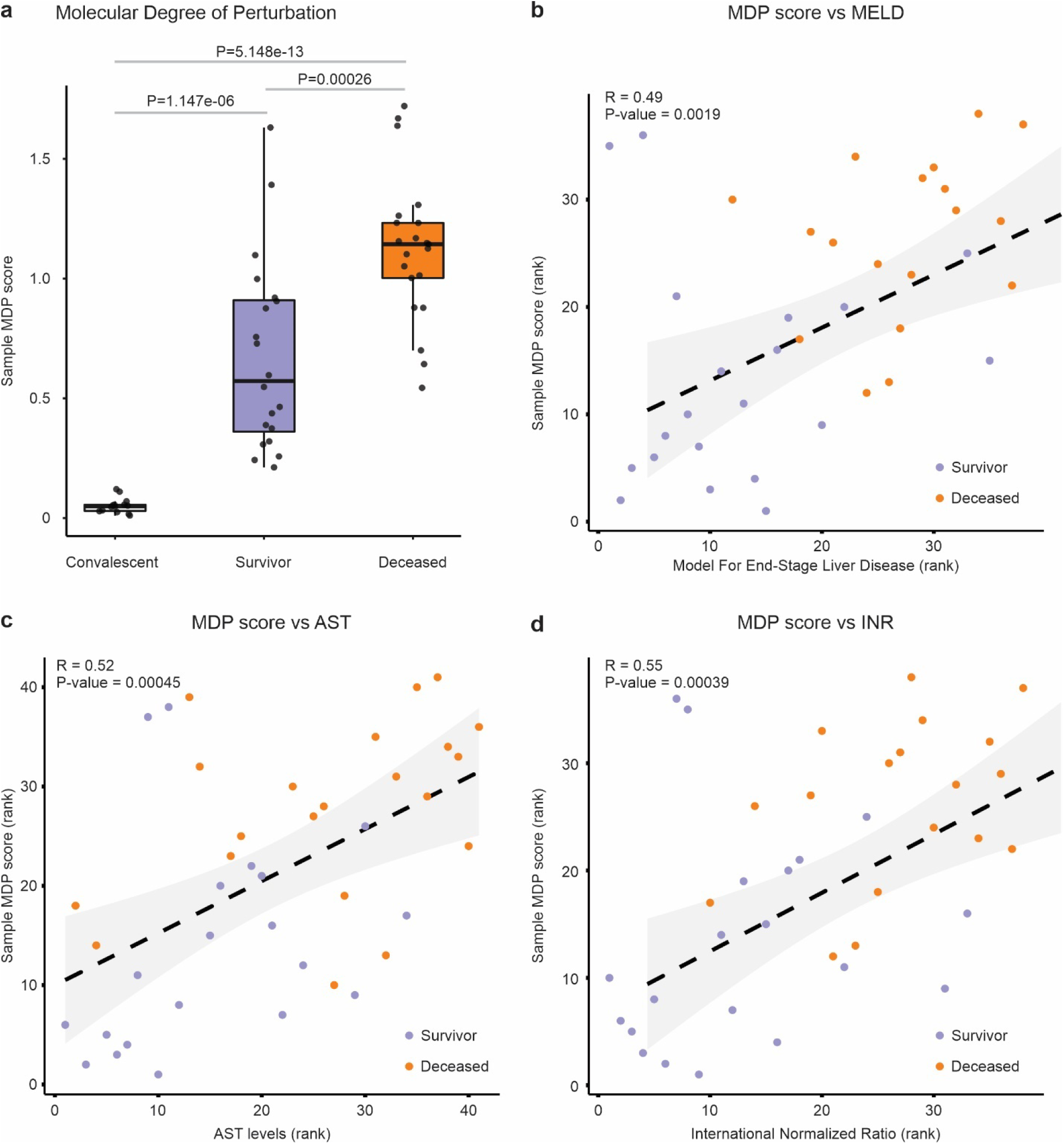
Assessing the Molecular Degree of Perturbation in Deceased and Surviving Patients. (a) The MDP score was calculated for each sample (black dots) using normalized expression values and outcome information, with the reference group consisting of patients in the convalescent phase. A nonparametric Wilcoxon test was used to assess sample MDP scores between patients in the convalescent phase and between deceased and surviving patients. Spearman correlation analysis comparing sample MDP score and (b) MELD score, (c) AST laboratory parameter, and (d) INR laboratory parameter. MDP (Molecular Degree of Perturbation); MELD (Model for End-Stage Liver Disease); AST (Aspartate Aminotransferase); INR (International Normalized Ratio).

**Figure S3.**
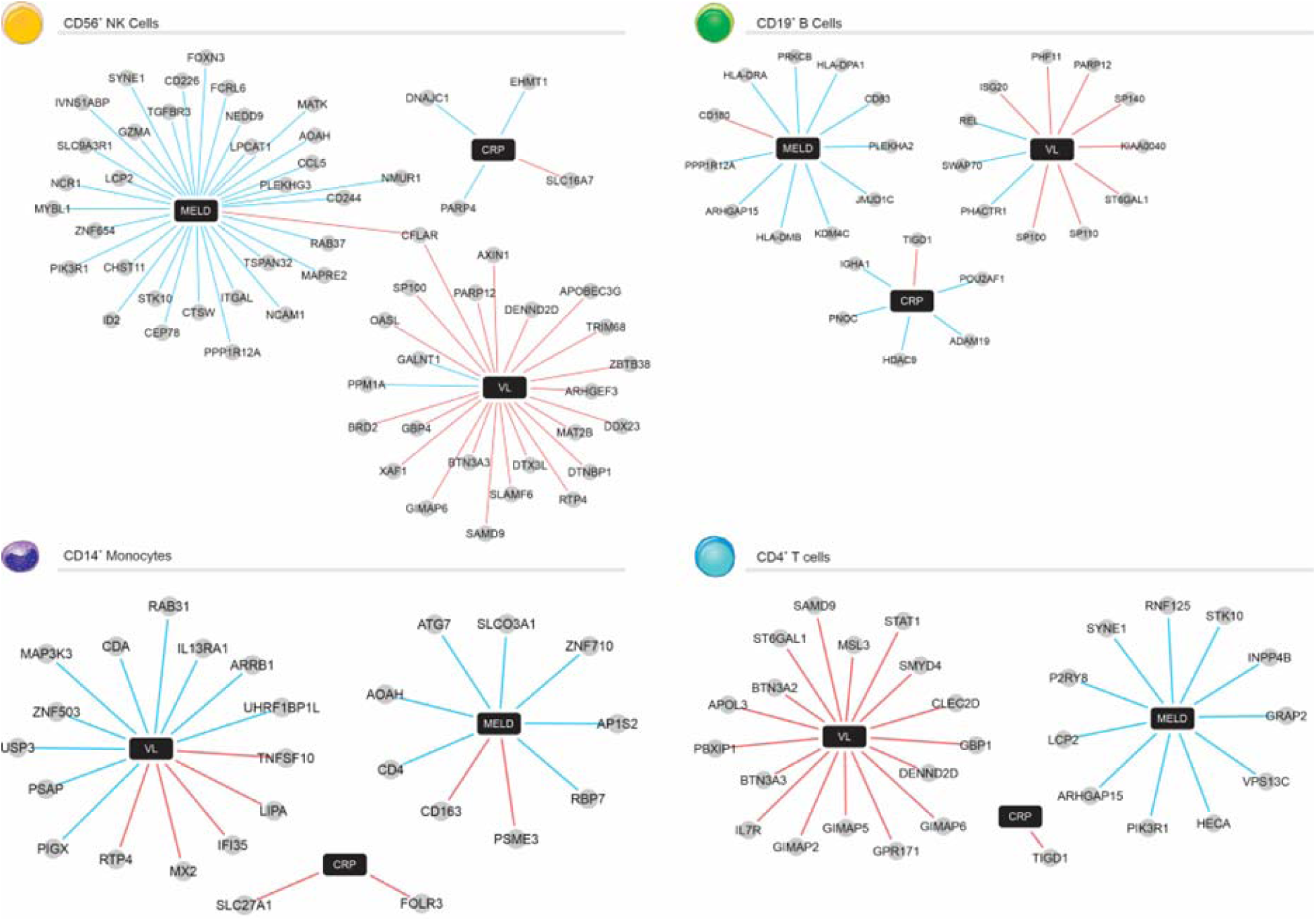
Signatures associated with Clinical Parameters in Immune Cell Populations. The network was constructed using the correlation between gene expression (normalized to patients in the convalescent phase) and MELD score, C-reactive protein (CRP), or viral load (VL) values. Correlated genes (Spearman’s rank correlation coefficient > 0.5) that are also expressed in Natural Killer cells, B cells, monocytes, or CD4^+^ T cells according to the Human Gene Atla were selected as nodes. The red line indicates a positive correlation, and the blue line indicates a negative correlation between genes (gray circles) and parameters (black rectangle).

**Figure S4.**
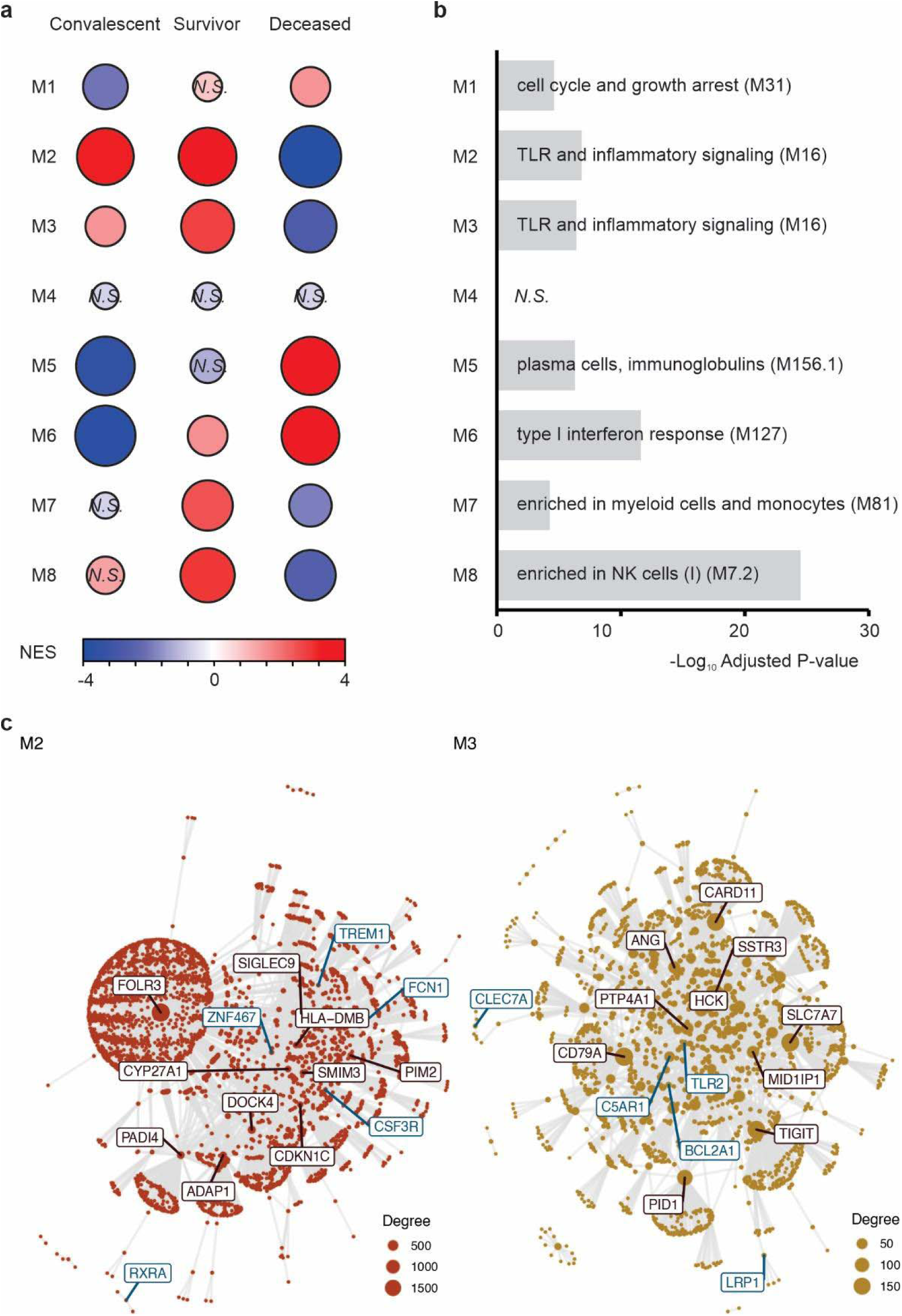
Modular Co-expression Analysis of YFV Infection. (a) Gene Set Enrichment Analysis (GSEA) of the 8 co-expression modules identified by CEMiTool in patients in the convalescent phase and in acute infection who either survived or died from the disease. The size and color are proportional to the Normalized Enrichment Score (NES). The rankings used in the GSEA analysis were obtained through z-score normalization per gene, followed by averaging the values across the 3 different groups. (b) Over-representation analysis between genes from each module and genes in the BTMs. An adjusted P-value cut-off of 0.05 was used. A representative BTM was selected for the bar chart. (c) Protein-protein interaction networks between genes in modules M2 and M3. Highly connected genes (hubs) are marked and were identified by the CEMiTool.

